# Birth outcomes in single motherhood ‘by choice’: A register study of children conceived through Medically Assisted Reproduction

**DOI:** 10.1101/2025.09.30.25336967

**Authors:** Alina Pelikh, Hanna Remes, Mine Kühn, Pekka Martikainen, Alice Goisis

**Affiliations:** Centre for Longitudinal Studies, Social Research Institute, University College London, 55-59 Gordon Square, London WC1E 6BT, United Kingdom; Helsinki Institute for Demography and Population Health, Faculty of Social Sciences, University of Helsinki, Finland; Max Planck – University of Helsinki Center for Social Inequalities in Population Health, Helsinki, Finland; Tilburg University, School of Social and Behavioral Sciences, Department of Sociology, PO Box 90153, 5000, LE Tilburg, Netherlands; Max Planck Institute for Demographic Research, Konrad-Zuse-Str. 1, 18055, Rostock, Germany

**Keywords:** birth outcomes, low birth weight, preterm birth, Medically Assisted Reproduction, single motherhood

## Abstract

**Background:** Our aim was to evaluate birth outcomes among children born to single mothers through Medically Assisted Reproduction (MAR), a rapidly growing and under studied group.

**Methods:** Using Finnish population register data from 1996-2020, we compared birth weight (grams); gestational age (days); low birth weight (LBW, <2500 grams), and preterm birth (<37 weeks)) of first children born to single mothers via MAR (n=2763) to outcomes of children born to single mothers who conceived spontanesously (SC; n=45 168) and children born to partnered mothers through MAR (n=38 693) or SC (n=413 326). We compared the outcomes before and after accounting for differences in maternal socio-demographic and health characteristics (maternal age, education, income, smoking, mental health), and multiplicity.

**Results:** Children conceived by single MAR mothers had, on average, a higher prevalence of LBW (8.0%) and prematurity (10.0%), compared to children of single SC mothers (5.7% and 6.2% respectively) and children of partnered NC mothers (4.3% and 5.5% respectively). However, their birth outcomes were generally better than those of children born to partnered MAR mothers (9.7% LBW, 11.8% premature). After accounting for differences in maternal socio-demographic and health characteristics, and multiplicity in a regression, differences between birth outcomes of single MAR mothers and partnered SC mothers fully attenuated, while children born to single SC and MAR partnered mothers had worse outcomes.

**Conclusions:** Single mothers who conceive via MAR are not at increased risk of poorer birth outcomes because of the MAR treatments or the single parenthood but rather because they have high rates of multiple births.

**Key Messages:**

- This is the first population-level study to investigate birth outcomes among children born to single mothers through Medically Assisted Reproduction (MAR), a rapidly growing and under studied group.
- Although unadjusted analyses suggested increased risks among children of single MAR mothers, results adjusted for socio-demographic characteristics and multiplicity showed their outcomes were, on average, better than those of children born to single women who conceived spontaneously (SC) as well as partnered MAR mothers, and similar to those of children born to partnered SC mothers.
- The results suggest that the advantaged characteristics of single MAR mothers offset the potential negative effects of both MAR treatments and single motherhood on birth outcomes.

## Introduction

Since the birth of the first IVF baby in 1978, over 10 million children worldwide have been conceived through assisted reproduction techniques (ART).^1^ As the use of infertility treatments increases globally, understanding its impact on children’s health outcomes is crucial. Studies on birth outcomes among children conceived through Medically Assisted Reproduction (MAR; including ovulation induction (OI), artificial insemination (AI) and ART) have found higher risks of adverse perinatal outcomes, such as low birth weight and preterm, compared to spontaneously conceived (SC) children.^2–5^ A growing subgroup within this population is children born to single mothers ‘by choice’ (women who conceive through MAR without a partner).^6–8^ Existing studies often only distinguish between married and unmarried women,^4,5^ but fail to look specifically at children born to single mothers through MAR. This is a significant knowledge gap not only because of the rapid increase of this group^9–12^ but also because there is evidence that children born to single mothers face increased risk of poorer birth outcomes,^13,14^ which can have long-term implications for children’s health, social outcomes, and public health costs.^15–17^ It is important to determine whether this is also true for single mothers who conceive through MAR.

Single MAR mothers share traits with both single SC mothers and partnered MAR mothers, but also possess characteristics distinguishing them from both group which makes it difficult to predict a priori what their children’s birth outcomes might look like. They may experience higher stress levels during conception and pregnancy due to lack of practical and emotional support of a partner or societal stigma associated with single motherhood^18,19^ (similar to single SC mothers) and stress associated with undergoing infertility treatments^20–24^ (similar to partnered MAR mothers), potentially negatively impacting birth outcomes. In addition, women who undergo MAR are on average older^2,4,5^ and tend to report poorer mental health prior to birth,^23^ factors that may further contribute to adverse birth outcomes.^4,5^ However, they often have advantageous socioeconomic characteristics similar to partnered MAR mothers, such as higher education and income levels,^25–29^ as well as better health behaviours^4,5^ (e.g., lower rates of pregnancy smoking) which could have protective effects. Two key factors further characterise single MAR mothers. First, they have strong fertility intentions and their pregnancies are intended, a factor strongly associated with better outcomes.^30^ Second, unlike partnered MAR mothers, single MAR mothers do not necessarily suffer from subfertility, which is found to be associated with worse birth outcomes.^2–5^ These unique characteristics of single MAR mothers represent an opportunity to learn about the impact of MAR and single motherhood on children’s birth outcomes.

This study utilises Finnish population register data to analyse birth outcomes of single MAR mothers in Finland from 1996 to 2020. This data contains a wide range of socio-economic and health indicators which allows to account for confounding in the associations with birth outcomes. To disentangle the potential effects of single parenthood and MAR treatments, we compare these outcomes to three other population groups: single SC mothers, partnered MAR mothers, and partnered SC mothers. Since 2006, single women in Finland have been granted access to infertility treatments in the public sector. However, in practice, before 2017, single women were unlikely to receive fertility treatments in the public sector as priority was given to patients with medical infertility, meaning that most single women underwent treatments in private clinics.

## Methods

### Study Population

We used birth records of all first-born children born in Finland between 1996 and 2020 (n=564 582). We focused only on first births to ensure comparable analyses across groups because MAR children are predominantly first-borns, ^2,4,5^ which is typically associated with worse birth outcomes compared to higher-order parities.^31^ We excluded births to women under 20 years of age (n=19 277) as undergoing MAR treatments before that age is rare. We further excluded quadruplets (n=4), stillbirths (n=669). We kept one record per first birth (n=535 765) in twin and triplet pregnancies and excluded cases with missing data on child’s birth weight or gestational age (n=1141). Finally, we excluded women whom we could not reliably identify as single mothers (n=5397) (discussed in detail in the next section and Supplementary Box S1). The final sample comprised 529 227 live births, with 41 670 (7.8%) conceived through MAR (Figure 1).

**Figure 1.**
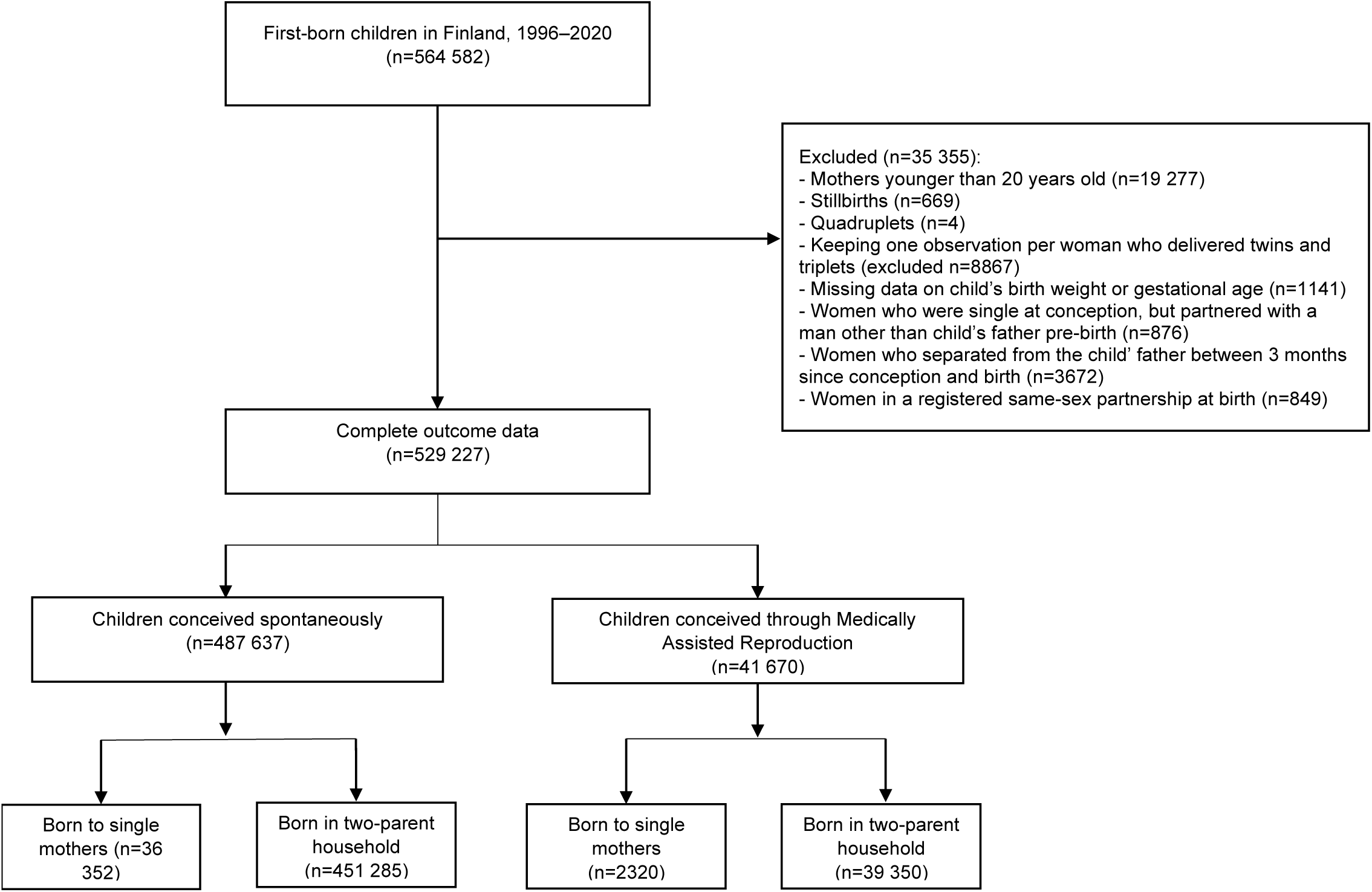
Study sample flow diagram

### Identifying single mothers ‘by choice’

We identified children born through MAR treatments using administrative data from the Birth Register, with MAR births recorded since 1996, complementing it with data on MAR treatments and purchases of infertility drugs. The Social Insurance Institution provides data on all individual reimbursement records, including purchases of infertility drugs used during MAR in public as well as private sector since 1995. For more details on this data, see Goisis et al. (2023).^23^ Additionally, the Institution provides data on MAR treatments conducted at private clinics, while the national Healthcare Register contains information on treatments performed at public clinics. We identified children conceived through MAR by combining each woman’s purchases of fertility drugs or the dates of MAR procedures with her child’s birthdate.

We identified single mothers using Statistics Finland’s cohabitation data, which includes information on the timing of moving in together, marriage, separation, divorce, and partner’s death for opposite-sex couples since 1987. A woman was classified as a single mother if she was not living with a partner between conception (estimated as birthdate minus gestational age) and birth, with some exceptions. Women who began cohabiting with the child’s father within six months post-birth were not classified as single mothers. As the cohabitation register does not contain information on same-sex couples and the mechanisms driving birth outcomes in these couples could be different,^32^ we excluded births to same-sex couples as reported in the Birth register (n=849). For further minor exclusions (n=4548) see Supplementary Table S1.

### Birth outcomes

We analysed four birth outcomes: birth weight (in grams) and gestational age (in days, calculated from available data on full weeks of gestation) (continuous), low birth weight (LBW, less than 2500 g), preterm birth (less than 37 weeks of gestation). Given the changes in prevalence, access, and regulations surrounding MAR in Finland over the 25-year observation period, we adjusted all models for the 5-year intervals (1996–2000, 2001–2005, 2006–2010, 2011–2015, 2016–2020).

### Covariates

First, we controlled for a set of variables related to maternal health and socio-economic status. We controlled for maternal age (20–24; 25–29; 30–34; 35–39; 40+), level of education (basic, secondary, tertiary), income (quintiles of the households’ equivalent disposable income pre-birth). We additionally included whether the mother smoked during pregnancy as an important risk-factor for children’s health.^33,34^ We adjusted for maternal mental health, as studies suggest that women undergoing MAR often have higher prevalence of mental ill-health pre-conception^23^ which may affect birth outcomes.^35–37^ To construct a measure of a woman’s mental health up to five years pre-MAR we used data on purchases of anxiolytics (ATC codes N05B), hypnotics and sedatives (N05C), and antidepressants (N06A), drugs often used to treat anxiety, depression, insomnia, and related mental health conditions. We grouped women as purchasers (≥1) and non-purchasers within this timeframe, considering only purchases from age 18 onwards. Second, we accounted for multiplicity (twins/triplets), which are more prevalent in MAR pregnancies due to treatment procedures and could mediate the association between MAR and birth outcomes.^2–5^

### Statistical analysis

We used linear models for continuous outcomes (birth weight and gestational age) and logistic models for binary outcomes (LBW and preterm birth), estimating three regressions for each outcome. Model 1 presents the baseline association between MAR and each of the outcomes controlling only for period and child’s sex. Model 2 introduces controls for maternal characteristics (maternal age at birth, level of education, income, smoking during pregnancy, history of psychotropic medication). Model 3 (fully adjusted) adds controls for multiplicity. For sensitivity analyses, we restricted the sample to singletons to remove the influence of multiple births and examined the period 2006–2020, when births to single MAR mothers were most common.

## Results

MAR treatments were utilised in 7.8% (n=41 670) of pregnancies; among MAR mothers, 5.4% conceived while single, versus 7.5% of SC mothers. The share of single mothers among first-time MAR conceptions rose to 10.2% in 2016-2020, while remaining stable at 7-8% for spontaneous conceptions (Figure 2).

**Figure 2.**
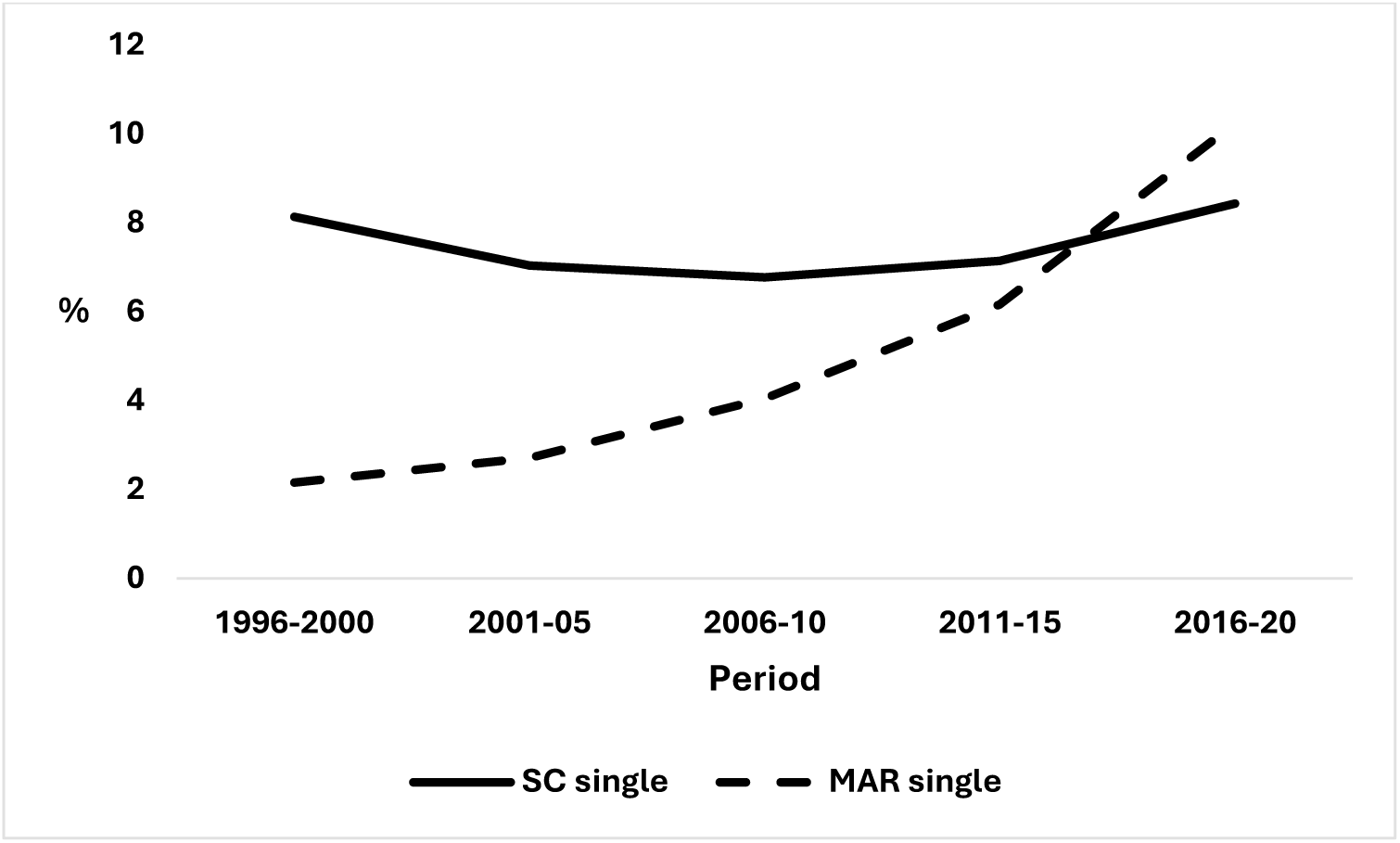
Proportion of single women among first-time SC and MAR mothers *Note*: ‘MAR’ refers to medically assisted reproduction, ‘SC’ stands for spontaneous conception. *Source*: Authors’ calculations from Finnish population register data.

Children born to single MAR mothers had, on average, lower birth weight and gestational age compared to SC children. They also had a higher proportion of low birth weight (LBW) and preterm births (Table 1). Among MAR children, those born to single mothers had, on average, better outcomes than those born to partnered mothers. In contrast, among SC children, those born to partnered mothers had the lowest prevalence of adverse birth outcomes.

**Table 1.** Descriptive statistics of birth outcomes of first-born children in Finland, 1996-2020, by mode of conception and maternal partnership status.

|  | MAR single | SC single | MAR partnered | SC partnered |
| --- | --- | --- | --- | --- |
|  | (n=2230) | (n=36 352) | (n=39 350) | (n=451 295) |
| **Birth outcomes** |  |  |  |  |
| Birth weight (grams) | 3353 | 3383 | 3313 | 3436 |
| 95% CI | (3326-3379) | (3377-3389) | (3306 - 3319) | (3434 - 3438) |
| Gestational age (weeks) | 39.0 | 39.4 | 38.8 | 39.4 |
| 95% CI | (38.9-39.1) | (39.3-39.4) | (38.8-38.8) | (39.4-39.4) |
| Low birth weight (%), <2500 grams | 8.0 | 5.7 | 9.7 | 4.3 |
| 95% CI | (6.7-8.9) | (5.5-5.9) | (9.4-10.0) | (4.3-4.4) |
| Preterm (%), <37 weeks | 10.0 | 6.2 | 11.8 | 5.5 |
| 95% CI | (8.3-10.7) | (6.0-6.5) | (11.5-12.1) | (5.4-5.6) |

*Note:* ‘MAR’ refers to medically assisted reproduction, ‘SC’ stands for spontaneous conception. 95% confidence intervals are presented underneath the mean statistics.
*Source:* Authors’ calculations from Finnish population register data.

The demographic profile of single MAR mothers differed notably from other groups of mothers (Table 2). On average, they were 3 years older than partnered MAR mothers and 7.5 years older than SC mothers. Education levels were highest among single MAR mothers (66.8% with tertiary education), followed closely by partnered MAR mothers (64.3%). Single SC women had the lowest education levels (22.7% with tertiary education). Income distribution for single MAR mothers was comparable to partnered SC mothers. Partnered MAR women had the highest average income, while single SC mothers had the lowest. Smoking during pregnancy was less prevalent among MAR mothers (6.9% single, 5.3% partnered), with single SC women reporting the highest proportion of smoking (35.2%). Single women, particularly those using MAR, were more likely to have used psychotropic medication within five years pre-conception (28.3% for single MAR mothers, 22.7% for single SC mothers) compared to partnered women in both groups. MAR-conceived children were more likely to be multiple births.

**Table 2.** Characteristics of women giving first birth in Finland, 1996-2020, and their children, by mode of conception and maternal partnership status.

|  | MAR<br>single | SC single | MAR<br>partnered | SC<br>partnered | All |
| --- | --- | --- | --- | --- | --- |
| Woman's age at birth (%) |  |  |  |  |  |
| 20–24 | 4.5 | 39.1 | 4.5 | 24.7 | 24.1 |
| 25–29 | 12.2 | 27.0 | 26.0 | 39.6 | 37.6 |
| 30–34 | 24.0 | 19.3 | 40.1 | 26.3 | 26.9 |
| 35–39 | 34.3 | 11.2 | 22.9 | 8.1 | 9.5 |
| 40+ | 24.9 | 3.4 | 6.5 | 1.3 | 1.9 |
| Woman's mean age at birth, (sd) | 35.7(5.7) | 27.9(5.9) | 32.5(4.7) | 28.5(4.7) | 28.8(4.9) |
| Period |  |  |  |  |  |
| 1996–2000 | 7.6 | 21.7 | 19.5 | 19.8 | 19.8 |
| 2001–2005 | 7.9 | 19.5 | 16.2 | 20.7 | 20.2 |
| 2006–2010 | 15.4 | 20 | 20.7 | 22.2 | 21.9 |
| 2011–2015 | 25.6 | 20 | 22.1 | 20.9 | 20.9 |
| 2016–2020 | 43.5 | 18.8 | 21.6 | 16.4 | 17.1 |
| Educational level |  |  |  |  |  |
| Basic | 8.1 | 31.6 | 5.6 | 12.6 | 13.4 |
| Secondary | 25.1 | 45.7 | 30.1 | 43.2 | 42.4 |
| Tertiary | 66.8 | 22.7 | 64.3 | 44.2 | 44.3 |
| Income quintile (year pre-birth) |  |  |  |  |  |
| 1st quintile | 9.1 | 33.1 | 4.0 | 13.8 | 14.4 |
| 2nd quintile | 10.0 | 18.4 | 5.2 | 12.2 | 12.1 |
| 3rd quintile | 16.6 | 16.8 | 9.4 | 15.2 | 14.8 |
| 4th quintile | 26.2 | 13.3 | 21.7 | 22.4 | 21.8 |
| 5th quintile | 33.0 | 9.9 | 56.3 | 30.0 | 30.6 |
| Missing | 5.0 | 8.6 | 3.3 | 6.3 | 6.3 |
| Smoking during pregnancy |  |  |  |  |  |
| No | 90.4 | 61.8 | 92.3 | 83.4 | 82.6 |
| Yes | 6.9 | 35.2 | 5.3 | 14.3 | 15.0 |
| Unknown | 2.7 | 3.1 | 2.3 | 2.3 | 2.4 |
| Psychotropic medicine purchases within five years pre-MAR (%) |  |  |  |  |  |
| Yes | 28.3 | 22.7 | 15.2 | 13.0 | 13.9 |
| Multiplicity (%) |  |  |  |  |  |
| Twins and triplets | 6.1 | 1.1 | 8.6 | 1.0 | 1.6 |
| Child's sex: female (%) | 50.7 | 48.9 | 48.9 | 48.8 | 48.8 |
| Total N | 2230 | 36 352 | 39 350 | 451 295 | 529 227 |

*Note:* 'MAR' refers to medically assisted reproduction, 'SC' stands for spontaneous conception. 'sd' stands for standard deviation.
*Source:* Authors' calculations from Finnish population register data.

Table 3 shows coefficients for all birth outcomes for children born to single MAR, single SC, partnered MAR and partnered SC mothers. The coefficients for the control variables included in Model 2 and Model 3 are presented in Supplementary Tables S1-S4. In Model 1, children born to single MAR mothers overall weighted less, had lower gestational age, and were at higher risk of being LBW and preterm than SC children born to both single and partnered mothers. However, their birth outcomes were generally better compared to children of MAR partnered mothers.

**Table 3.** Models of birth outcomes (95% CI) of first-born children in Finland, 1996-2020, by mode of conception and maternal partnership status: a) birth weight; b) gestational age; c) low birth weight; d) preterm.

|  | Model 1 (baseline) | Model 2 (Model 1 +<br>maternal age + level of<br>education + income +<br>smoking during<br>pregnancy +<br>psychotropic<br>medication use five<br>years pre-MAR) | Model 3 (Model 2 +<br>multiple birth) |
| --- | --- | --- | --- |
| a) Birth weight (in grams). Coefficients from linear models |  |  |  |
| SC single (Ref: MAR single) | 30 (7,54) | 13 (-10, 37) | -39 (-62, -16) |
| MAR partnered | -40 (-63, -17) | -65 (-89, -42) | -42 (-65, -19) |
| SC partnered | 84 (61, 107) | 44 (21, 67) | -9 (-31, 13) |
| b) Gestational age (in days). Coefficients from linear models |  |  |  |
| SC single (Ref: MAR single) | 3.0 (2.4, 3.6) | 2.7 (2.2, 3.3) | 1.1 (0.6, 1.7) |
| MAR partnered | -1.1 (-1.6, -0.5) | -1.4 (-2.0, -0.9) | -0.9 (-1.4, -0.3) |
| SC partnered | 3.4 (2.8, 3.9) | 2.7 (2.2, 3.3) | 1.4 (0.8, 1.9) |
| c) Low birth weight. Odds ratios from logistic models |  |  |  |
| SC single (Ref: MAR single) | 0.69 (0.59, 0.81) | 0.77 (0.66, 0.91) | 1.21 (1.01, 1.45) |
| MAR partnered | 1.24 (1.05, 1.45) | 1.42 (1.21, 1.66) | 1.30 (1.08, 1.55) |
| SC partnered | 0.52 (0.44, 0.60) | 0.65 (0.55, 0.76) | 1.01 (0.85, 1.21) |
| d) Preterm. Odds ratios from logistic models |  |  |  |
| SC single (Ref: MAR single) | 0.61 (0.52, 0.71) | 0.69 (0.59, 0.80) | 0.96 (0.82, 1.13) |
| MAR partnered | 1.23 (1.07, 1.43) | 1.32 (1.14, 1.53) | 1.21 (1.04, 1.42) |
| SC partnered | 0.53 (0.46, 0.61) | 0.61 (0.53, 0.71) | 0.86 (0.74, 1.01) |

*Note:* 'MAR' refers to medically assisted reproduction, 'SC' stands for spontaneous conception. 95% CI are presented in parentheses. Baseline models control for period and child's sex.
*Source:* Authors' calculations from Finnish population register data.

In Model 2, after accounting for maternal socio-demographic and health characteristics, the differences in outcomes between MAR children born to single mothers and SC children became smaller. For example, the OR for being born LBW among children of single SC mothers increased from 0.69 (95%CI 0.59-0.81) to 0.77 (95%CI 0.65-0.91), among children of partnered SC mothers OR increased from 0.51 (95%CI 0.44-0.60) to 0.64 (95%CI 0.55-0.75) in comparison to children of single MAR mothers (reference category in the analyses). This is related to the fact that single MAR mothers had more advantaged socio-economic characteristics which were associated with better perinatal outcomes for children. In contrast, the OR of LBW among children of partnered MAR mothers increased from 1.24 (95%CI 1.05-1.45) in Model 1 to 1.41 (95%CI 1.21-1.66) in Model 2 compared to children of single MAR mothers.

After the final adjustment for multiple birth, the differences in birthweight, gestational age, OR for LBW and preterm between children of single MAR mothers and partnered SC mothers became small, with coefficients’ confidence intervals overlapping at 95% level. For example, the OR for LBW increased to 1.01 (95%CI 0.85-1.21). In contrast, the differences in outcomes compared to children of single SC mothers became larger in magnitude and reverse in the direction of the association, i.e., became worse for children of single SC mothers compared to children of single MAR mothers (with the exception for the preterm OR with overlapping 95% CI). For example, the OR for LBW increased to 1.21 (95%CI 1.01-1.45). The differences in magnitude of the outcomes for children of partnered MAR mothers became smaller, yet remained pronounced. For example, the OR for being born LBW decreased to 1.30 (95%CI 1.08-1.55). The results of the main analysis were highly similar if we excluded multiple births from the analytical sample (Supplementary Table S5). The OR for being born LBW and preterm for Model 1 (unadjusted baseline model) and Model 3 (fully adjusted for maternal and child’s characteristics) are presented in Figure 3.

**Figure 3.**
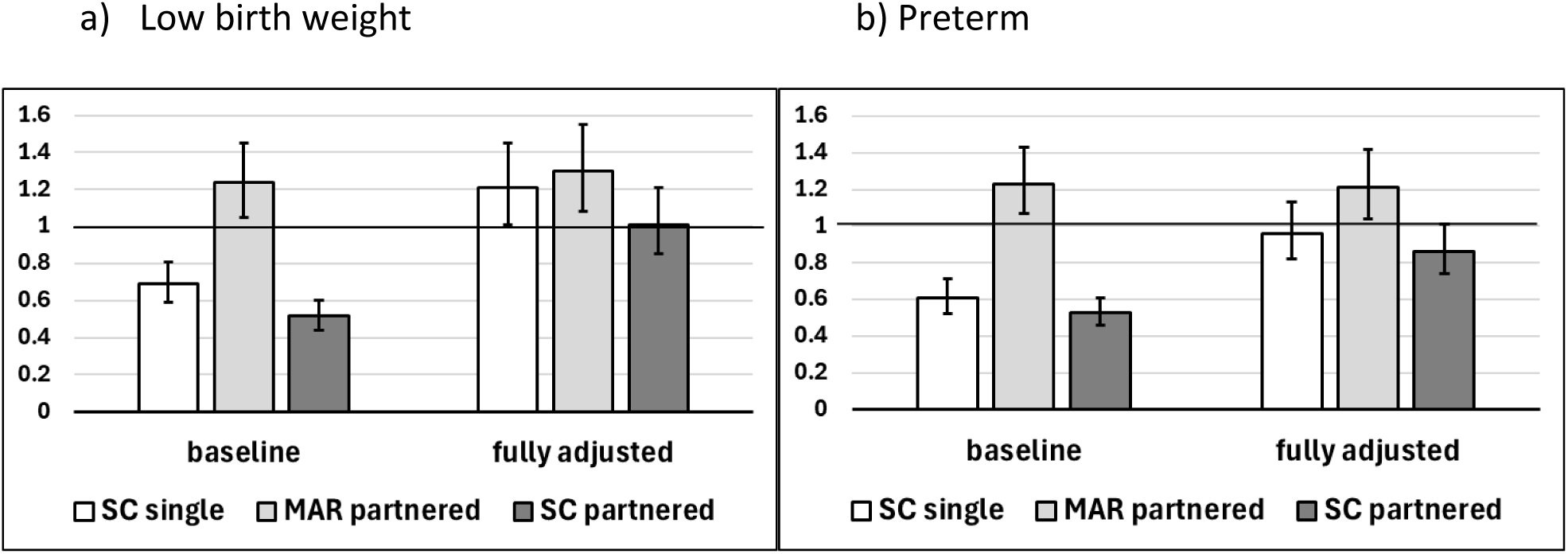
Odds ratios and 95% confidence intervals for a) low birth weight; b) preterm birth for first-born children in Finland, 1996-2020, by mode of conception and maternal partnership status *Note*: ‘MAR’ refers to medically assisted reproduction, ‘SC’ stands for spontaneous conception. The solid line represents single mothers who conceived via MAR, the reference category in the models(=1). Baseline model controls only for period and child’s sex and presents the unadjusted descriptive association between MAR and low birth weight (a) and preterm birth (b). Fully adjusted models are controlled for maternal age, level of education, income, smoking during pregnancy, psychotropic medication use five years pre-MAR, and multiple birth. *Source*: Authors’ calculations from Finnish population register data.

Additional sensitivity analyses (model 3 specification) excluding the time periods before 2006 (Supplementary Talbes S1-S4, column d), when treatments were prohibited to single women in the public sector, are in line with the main analyses. Hovewer, the relative advantage (e.g., differences in OR for LBW, Supplementary Table S3)) of children of single MAR mothers in comparison to those of single SC and partnered MAR mothers was more pronounced in these analyses.

## Discussion

### Main findings and contributions

Using Finnish register data, we investigated birth weight, gestational age, the risk of LBW and being born preterm among children of single MAR mothers. In the baseline models, they had, on average, a higher prevalence of LBW and prematurity compared to children of single SC and partnered SC mothers. In contrast, their birth outcomes were generally better than those of children of partnered MAR mothers. After accounting for maternal socio-demographic and health characteristics and multiplicity, differences between birth outcomes of single MAR and partnered SC mothers fully attenuated, while children born to single SC and partnered MAR mothers had worse outcomes. Three main implications emerge from the results.

First, the high prevalence of multiple births among MAR-conceived children are likely to be the primary driver of the increased risk of poor birth outcomes observed in the unadjusted analyses amongst children of single MAR mothers. The analyses including only singleton births confirmed these conclusions. Notably, our analyses focusing on more recent years (2006-2020), during which single women had wider access to MAR treatments, suggest that these regulatory changes were associated with improved birth outcomes for their children. One of the drivers behind this improvement could be the substantial decrease in the proportion of multiple births among single MAR mothers from 12.4% in 1996-2000 to 3.6% in 2016-2020 (Supplementary Table S6).

Second, MAR treatments per se are unlikely to play a major role in determining birth outcomes, as adjusted results show children of single MAR mothers have similar outcomes to those of partnered SC mothers and better outcomes than those of partnered MAR mothers. The fact that children of single MAR mothers have better birth outcomes than those of partnered MAR mothers likely reflects that only the latter suffer from subfertility, a risk factor for poor birth outcomes. This aligns with research showing that children of lesbian couples have similar birth outcomes to children conceived spontaneously,^32^ as both lesbian couples and single MAR mothers undergo MAR to conceive but typically not because they are subfertile.

Third, these results challenge assumptions about single motherhood by showing that when being a single mother goes together with high socioeconomic status and pregnancy planning – like in the case of single MAR mothers – is not associated with poorer outcomes for children. There are two potential interpretations: one is that it is not single motherhood per se that leads to poorer outcomes for children, it is the disadvantages that normally accompany growing up with a single parent. The other is that the selected and advantaged characteristics of single MAR mothers might offset the negative effects associated with single parenthood.

To best of our knowledge, this is the first population-level investigation of birth outcomes among children of single MAR mothers or single mothers ‘by choice’, a rapidly growing group. This research is timely given delayed childbearing trends and evolving treatment access regulations. As partnership trajectories preceding parenthood become more complex,^29^ more women may pursue solo motherhood. It is important to note that many of the single mothers ‘by choice’ have reported that they would have liked to be in a relationship instead of pursuing MAR alone, but they could not afford to wait any longer for the suitable partner.^8,25,39,40^ Our study examined a context where MAR treatments are largely publicly subsidized, resulting in a more diverse population of women accessing these services compared to countries with predominantly private funding. However, we believe the observed associations may be generalizable to other contexts as social acceptance grows and single women’s treatment access expands globally.^41^

While the rise in single motherhood has often been linked to lower socioeconomic status and negative outcomes for children,^9–12^ this pattern may not apply to single MAR mothers. These women typically have strong fertility intentions and sufficient resources to raise a child independently, partly distinguishing them from single mothers who conceive spontaneously. For birth outcomes, these factors appear to offset the challenges from potential stress and stigma of both single parenthood and MAR treatment. By examining birth outcomes in this specific population subgroup at a national level, our study contributes to the literature on children’s outcomes of single MAR mothers. While existing research suggests no long-term negative effects on children’s psychological wellbeing or quality of mother-child relationship,^42–44^ these studies typically use small samples, highlighting the need for more population-level evidence.

### Strengths and limitations

Our study’s strength include the use of high-quality population register data that allows to compare birth outcomes of children of single MAR mothers to those of single SC mothers, and partnered MAR and SC mothers. These comprehensive population-level data avoid nonresponse and recall biases of survey data. We employed all available data sources to comprehensively identify children conceived through MAR, determine maternal partnership status at conception/birth, and capture maternal sociodemographic characteristics and mental health.

Our study has a few limitations. As cohabitation register does not allow to identify same-sex or living-apart-together couples, it is possible that some women classified as single had partners. Nevertheless, we believe these couples only represent few cases and will not affect the overall results from the analyses as we excluded identifiable same-sex couples and prevalence of living-apart-together couples is low among new parents as such partnerships tend to either dissolve or transition to cohabitation shortly after conception.^45–47^ To further minimise misclassification, we considered women who started living with the child’s father within six months post-birth as partnered.^46^ Our data could not differentiate treatment types (e.g., OI/AI/IVF) which are known to affect birth outcomes.^2,5^ However, due to potential underreporting of less invasive treatments in our data and single women’s greater likelihood of undergoing IVF,^8^ our sample likely overrepresents more invasive treatments, making our estimates comparing birth outcomes among single and partnered MAR women conservative.

## Conclusions

After accounting for multiplicity, birth outcomes of children born to single MAR mothers were, on average, better compared to those of children born to single SC women as well as partnered MAR mothers, and similar to those of children born to partnered SC mothers. This suggests that the advantaged characteristics of single MAR mothers offset the potential negative effects of both MAR treatments and single motherhood on birth outcomes.

## Ethics approval

This study was approved by the Ethics Committee of Statistics Finland’s permission TK-52-1121-18.

## Author contributions

All authors defined the research question and designed the study. A.P. performed the statistical analyses (data cleaning and calculations) and wrote the first draft. All authors critically contributed to the interpretation of the results, revised the manuscript, and approved the final version for publication.

## Use of artificial intelligence (AI) tools

ChatGPT was used to improve the readability and grammar. No AI tools were used for data analysis, interpretation of results, or writing of original scientific content. The authors take full responsibility for the content of the study.

## Data Availability

The data that support the findings of this study are available from Statistic Finland. Restrictions apply to the availability of these data, which were used under license for this study.

## Supplementary material

### Supplementary Box S1.

#### Further detail on identification of single mothers in the Finnish register data

We also excluded a small number of women who were single at conception but were living with a male partner other than the child’s father (n=876) at the time of birth as it is difficult to categorise these women as single or partnered. In cases where the mother separated from the father within 3 months post-conception and remained single until birth (n=2695, predominantly SC (97%)), we identified them as single as it is likely that the actual separation occurred earlier. For separations occurring between 3 months of conception and birth (n=3672, 97% SC), we excluded those women from the analysis as separation at later stages of pregnancy could negatively affect birth outcomes^48^ complicating the comparison to women who were single at conception and decided to have a child on their own (both MAR and SC).

---

**Supplementary Table S1.**
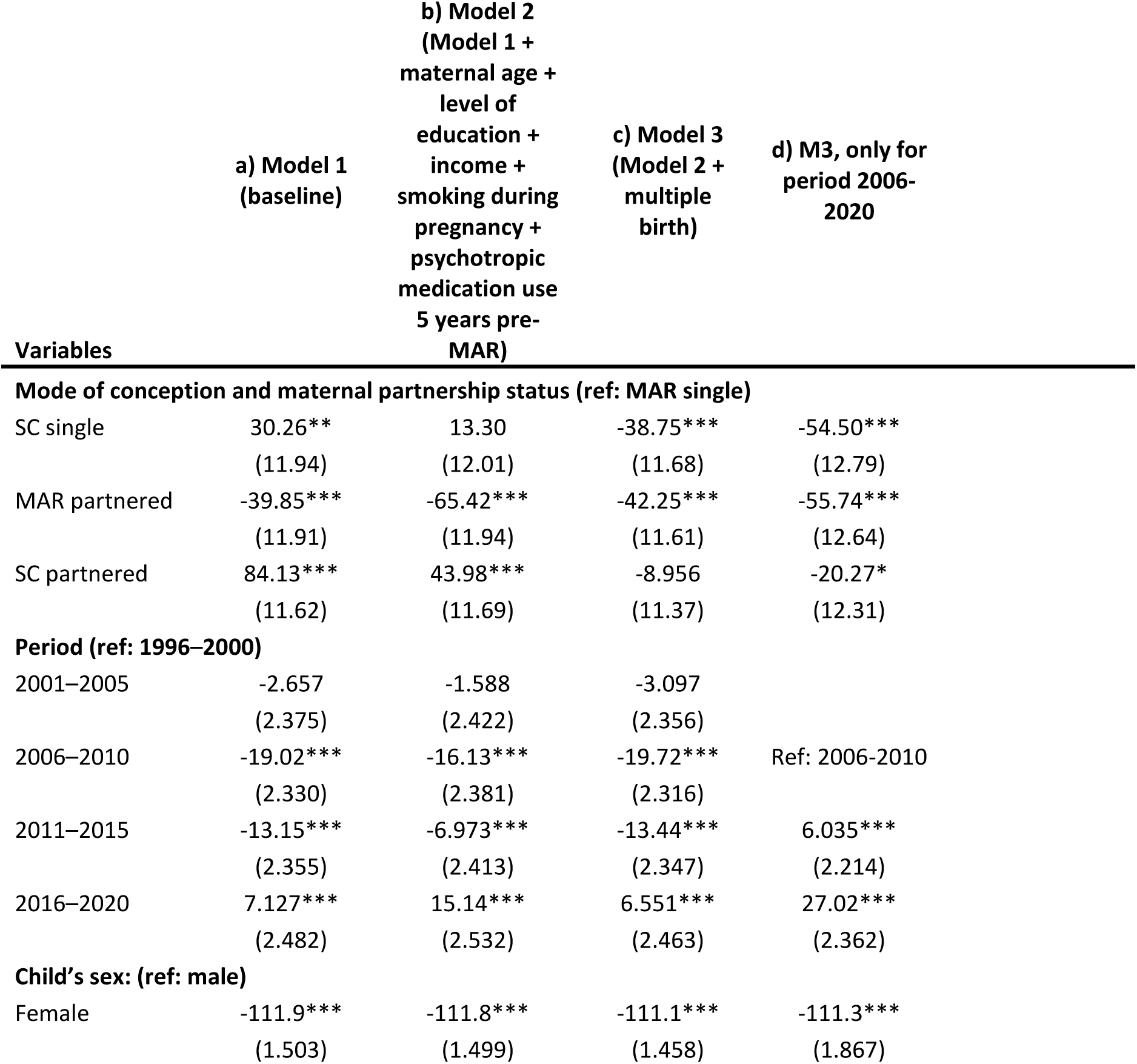
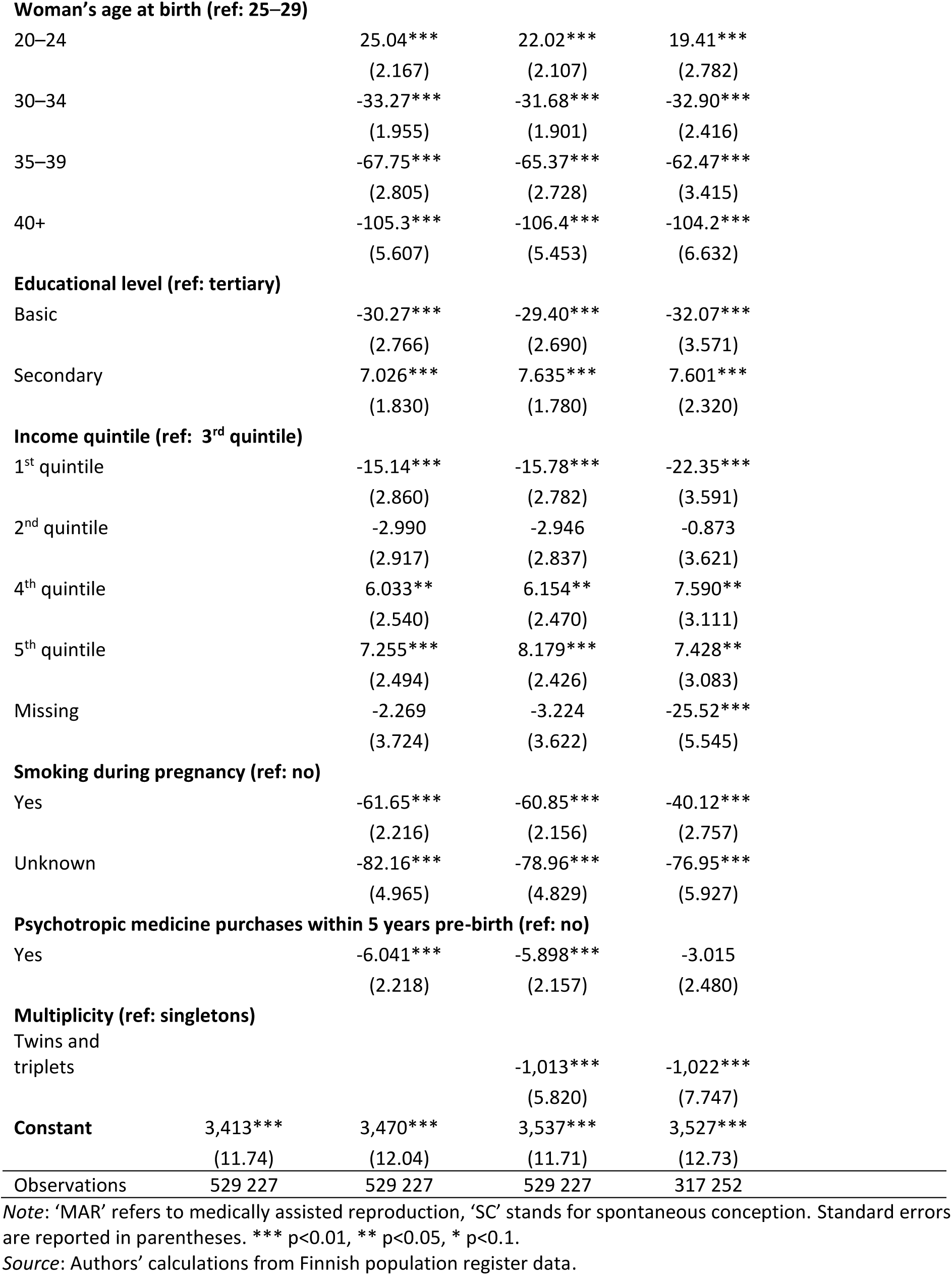
Linear models on birth weight (in grams) of first-born children born in Finland, 1996-2020.

**Supplementary Table S2.** Linear models on gestational age (in days) of first-born children born in Finland, 1996-2020.

| Variables | a) Model 1<br>(baseline) | b) Model 2<br>(Model 1 +<br>maternal age +<br>level of<br>education +<br>income +<br>smoking during<br>pregnancy +<br>psychotropic<br>medication use<br>5 years pre-MAR) | c) Model 3<br>(Model 2 +<br>multiple<br>birth) | d) M3, only for<br>period 2006-2020 |
| --- | --- | --- | --- | --- |
| **Mode of conception and maternal partnership status (ref: MAR single)** |  |  |  |  |
| SC single | 2.985***<br>(0.289) | 2.463***<br>(0.291) | 1.120***<br>(0.282) | 0.741**<br>(0.310) |
| MAR partnered | -1.077***<br>(0.288) | -1.450***<br>(0.290) | -0.852***<br>(0.281) | -1.197***<br>(0.306) |
| SC partnered | 3.366***<br>(0.281) | 2.748***<br>(0.284) | 1.382***<br>(0.275) | 1.074***<br>(0.298) |
| **Period (ref: 1996–2000)** |  |  |  |  |
| 2001–2005 | 0.433***<br>(0.0575) | 0.442***<br>(0.0588) | 0.403***<br>(0.0569) |  |
| 2006–2010 | 0.869***<br>(0.0564) | 0.919***<br>(0.0578) | 0.826***<br>(0.0560) | Ref: 2006-2010 |
| 2011–2015 | 0.853***<br>(0.0570) | 0.952***<br>(0.0585) | 0.785***<br>(0.0567) | -0.0495<br>(0.0536) |
| 2016–2020 | 0.916***<br>(0.0601) | 1.113***<br>(0.0614) | 0.892***<br>(0.0595) | 0.0415<br>(0.0571) |
| **Child's sex: (ref: male)** |  |  |  |  |
| Female | 0.672***<br>(0.0364) | 0.675***<br>(0.0364) | 0.694***<br>(0.0352) | 0.698***<br>(0.0452) |
| **Woman's age at birth (ref: 25–29)** |  |  |  |  |
| 20–24 |  | 0.387***<br>(0.0526) | 0.310***<br>(0.0509) | 0.188***<br>(0.0673) |
| 30–34 |  | -0.186***<br>(0.0474) | -0.145***<br>(0.0459) | -0.0959<br>(0.0584) |
| 35–39 |  | -0.598***<br>(0.0680) | -0.536***<br>(0.0659) | -0.429***<br>(0.0826) |
| 40+ |  | -1.585***<br>(0.136) | -1.614***<br>(0.132) | -1.452***<br>(0.160) |
| **Educational level (ref: tertiary)** |  |  |  |  |
| Basic |  | -0.222***<br>(0.0671) | -0.200***<br>(0.0650) | -0.335***<br>(0.0864) |
| Secondary |  | -0.148***<br>(0.0444) | -0.132***<br>(0.0430) | -0.228***<br>(0.0561) |
| **Income quintile (ref: 3<sup>rd</sup> quintile)** |  |  |  |  |
| 1<sup>st</sup> quintile |  | 0.174** | 0.158** | 0.108 |
|  |  | (0.0694) | (0.0672) | (0.0869) |
| 2<sup>nd</sup> quintile |  | 0.0738 | 0.0750 | 0.0453 |
|  |  | (0.0708) | (0.0686) | (0.0876) |
| 4<sup>th</sup> quintile |  | -0.0195 | -0.0163 | -0.0301 |
|  |  | (0.0616) | (0.0597) | (0.0753) |
| 5<sup>th</sup> quintile |  | 0.0228 | 0.0466 | 0.0492 |
|  |  | (0.0605) | (0.0586) | (0.0746) |
| Missing |  | -0.146 | -0.171* | -0.465*** |
|  |  | (0.0903) | (0.0875) | (0.134) |
| **Smoking during pregnancy (ref: no)** |  |  |  |  |
| Yes |  | -0.108** | -0.0874* | -0.123* |
|  |  | (0.0538) | (0.0521) | (0.0667) |
| Unknown |  | -2.385*** | -2.303*** | -2.185*** |
|  |  | (0.120) | (0.117) | (0.143) |
| **Psychotropic medicine purchases within 5 years pre-birth (ref: no)** |  |  |  |  |
| Yes |  | -0.618*** | -0.615*** | -0.547*** |
|  |  | (0.0538) | (0.0521) | (0.0600) |
| **Multiplicity (ref: singletons)** |  |  |  |  |
| Twins and triplets |  |  | -26.12*** | -27.05*** |
|  |  |  | (0.141) | (0.187) |
| **Constant** | 274.5*** | 275.2*** | 277.0*** | 278.2*** |
|  | (0.284) | (0.292) | (0.283) | (0.308) |
| Observations | 529 227 | 529 227 | 529 227 | 317 252 |

*Note:* 'MAR' refers to medically assisted reproduction, 'SC' stands for spontaneous conception. Standard errors are reported in parentheses. \*\*\* p<0.01, \*\* p<0.05, \* p<0.1.
*Source:* Authors' calculations from Finnish population register data.

**Supplementary Table S3.**
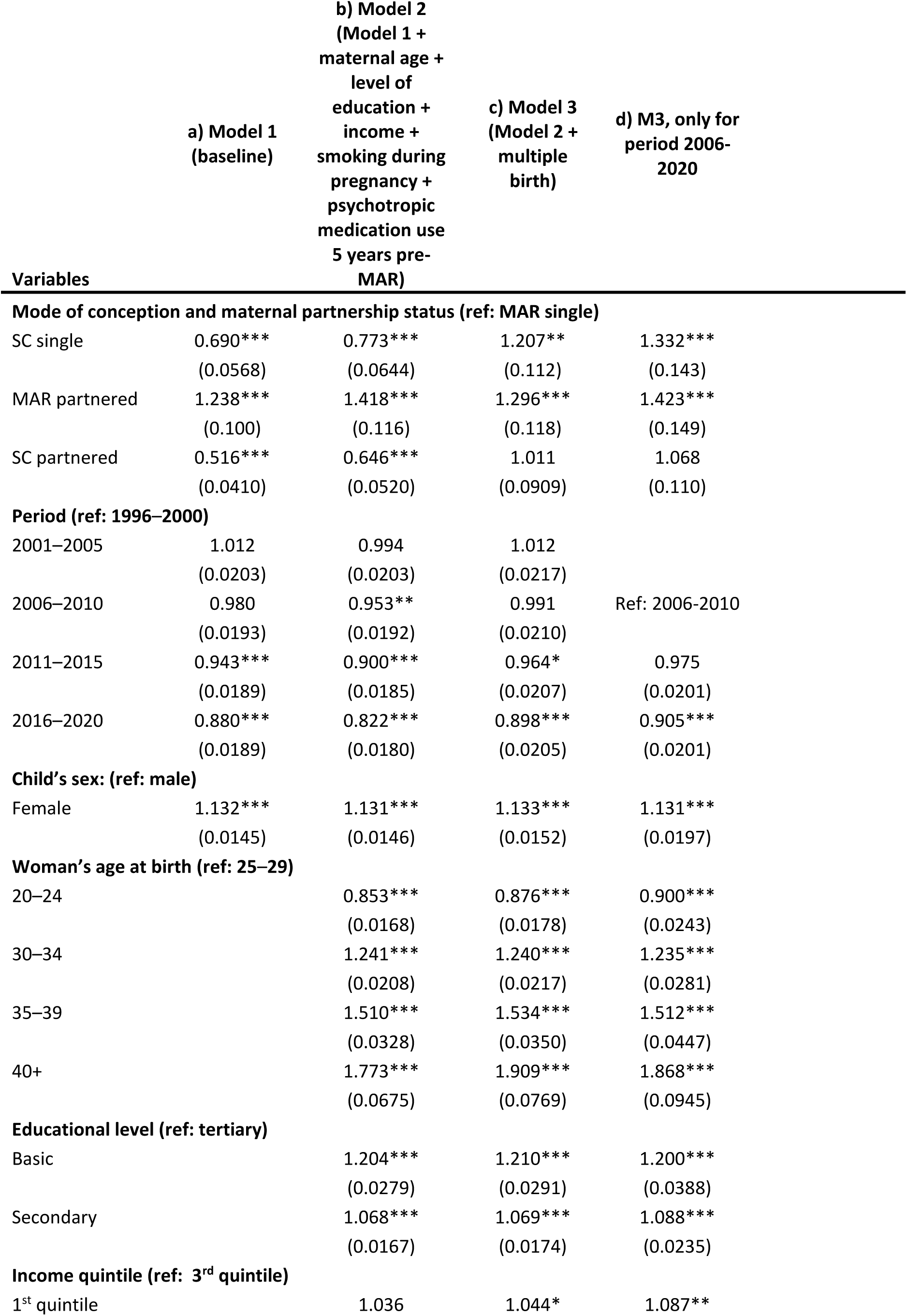
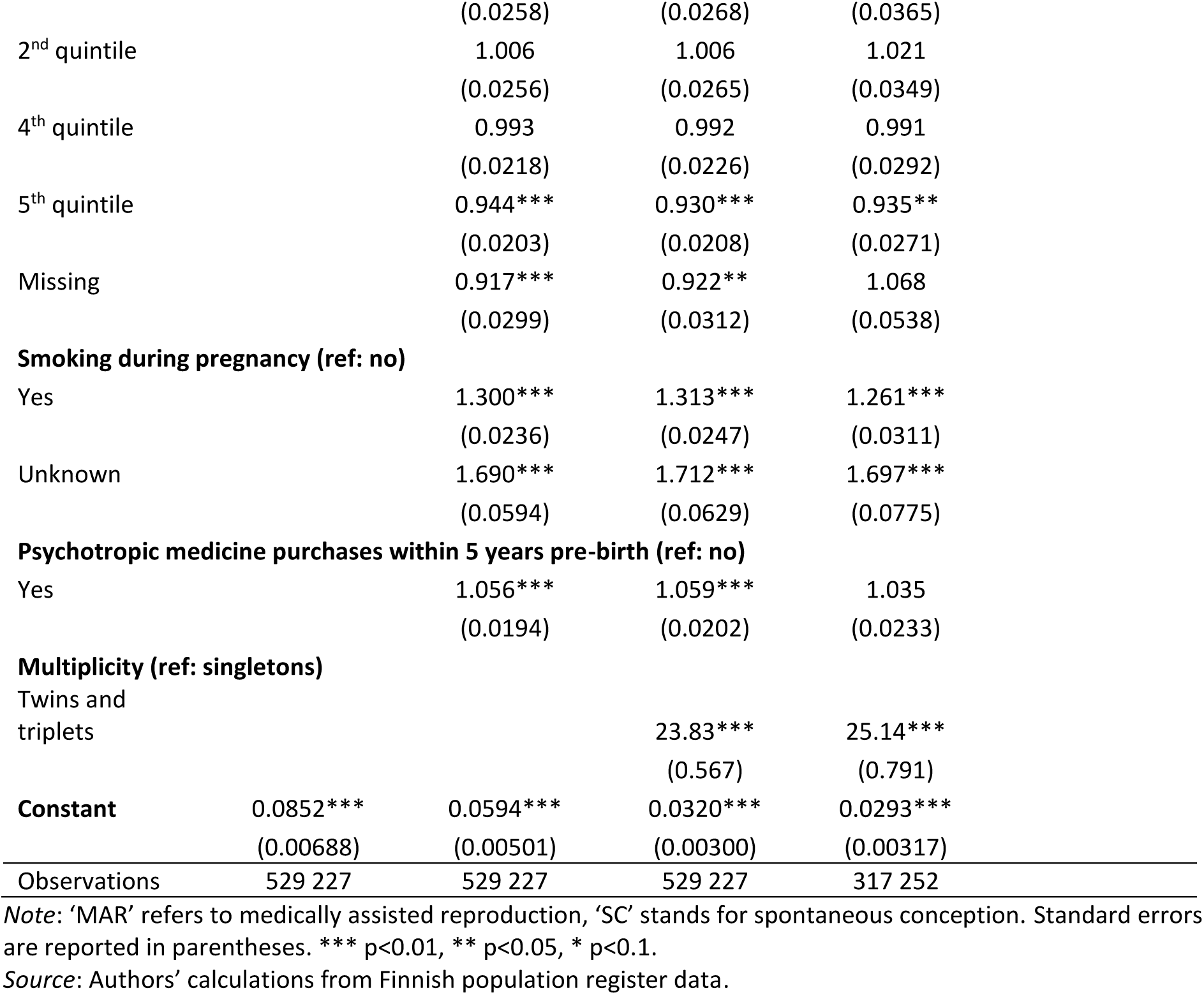
Odds ratios from logistic models on low birth weight of first-born children born in Finland, 1996-2020.

**Supplementary Table S4.** Odds ratios from logistic models on preterm of first-born children born in Finland, 1996-2020.

| Variables | a) Model 1<br>(baseline) | b) Model 2<br>(Model 1 +<br>maternal age +<br>level of<br>education +<br>income +<br>smoking during<br>pregnancy +<br>psychotropic<br>medication use<br>5 years pre-MAR) | c) Model 3<br>(Model 2 +<br>multiple<br>birth) | d) M3, only for<br>period 2006-2020 |
| --- | --- | --- | --- | --- |
| **Mode of conception and maternal partnership status (ref: MAR single)** |  |  |  |  |
| SC single | 0.610***<br>(0.0461) | 0.687***<br>(0.0525) | 0.963<br>(0.0799) | 1.032<br>(0.0995) |
| MAR partnered | 1.237***<br>(0.0915) | 1.322***<br>(0.0985) | 1.214**<br>(0.0988) | 1.332***<br>(0.125) |
| SC partnered | 0.534***<br>(0.0388) | 0.615***<br>(0.0452) | 0.864*<br>(0.0693) | 0.935<br>(0.0862) |
| **Period (ref: 1996–2000)** |  |  |  |  |
| 2001–2005 | 0.973<br>(0.0175) | 0.954**<br>(0.0175) | 0.965*<br>(0.0184) |  |
| 2006–2010 | 0.920***<br>(0.0163) | 0.896***<br>(0.0163) | 0.920***<br>(0.0173) | Ref: 2006-2010 |
| 2011–2015 | 0.913***<br>(0.0164) | 0.879***<br>(0.0162) | 0.927***<br>(0.0177) | 1.008<br>(0.0186) |
| 2016–2020 | 0.859***<br>(0.0165) | 0.809***<br>(0.0158) | 0.866***<br>(0.0175) | 0.941***<br>(0.0187) |
| **Child's sex: (ref: male)** |  |  |  |  |
| Female | 0.855***<br>(0.00995) | 0.855***<br>(0.00995) | 0.841***<br>(0.0101) | 0.836***<br>(0.0131) |
| **Woman's age at birth (ref: 25–29)** |  |  |  |  |
| 20–24 |  | 0.882***<br>(0.0155) | 0.902***<br>(0.0162) | 0.921***<br>(0.0222) |
| 30–34 |  | 1.115***<br>(0.0167) | 1.107***<br>(0.0171) | 1.110***<br>(0.0225) |
| 35–39 |  | 1.255***<br>(0.0252) | 1.254***<br>(0.0262) | 1.244***<br>(0.0336) |
| 40+ |  | 1.360***<br>(0.0500) | 1.409***<br>(0.0543) | 1.433***<br>(0.0688) |
| **Educational level (ref: tertiary)** |  |  |  |  |
| Basic |  | 1.087***<br>(0.0233) | 1.086***<br>(0.0239) | 1.085***<br>(0.0324) |
| Secondary |  | 1.054***<br>(0.0147) | 1.053***<br>(0.0152) | 1.076***<br>(0.0208) |
| **Income quintile (ref: 3<sup>rd</sup> quintile)** |  |  |  |  |
| 1<sup>st</sup> quintile |  | 0.951** | 0.954** | 0.962 |
|  |  | (0.0217) | (0.0223) | (0.0296) |
| 2<sup>nd</sup> quintile |  | 1.008 | 1.008 | 1.012 |
|  |  | (0.0231) | (0.0237) | (0.0309) |
| 4<sup>th</sup> quintile |  | 1.006 | 1.005 | 0.987 |
|  |  | (0.0198) | (0.0204) | (0.0259) |
| 5<sup>th</sup> quintile |  | 0.991 | 0.983 | 0.974 |
|  |  | (0.0191) | (0.0195) | (0.0251) |
| Missing |  | 0.913*** | 0.914*** | 1.061 |
|  |  | (0.0269) | (0.0277) | (0.0485) |
| **Smoking during pregnancy (ref: no)** |  |  |  |  |
| Yes |  | 1.066*** | 1.063*** | 1.095*** |
|  |  | (0.0184) | (0.0189) | (0.0252) |
| Unknown |  | 1.623*** | 1.637*** | 1.639*** |
|  |  | (0.0520) | (0.0542) | (0.0679) |
| **Psychotropic medicine purchases within 5 years pre-birth (ref: no)** |  |  |  |  |
| Yes |  | 1.076*** | 1.080*** | 1.065*** |
|  |  | (0.0179) | (0.0186) | (0.0216) |
| **Multiplicity (ref: singletons)** |  |  |  |  |
| Twins and triplets |  |  | 17.84*** | 19.87*** |
|  |  |  | (0.416) | (0.616) |
| **Constant** | 0.117*** | 0.0962*** | 0.0657*** | 0.0547*** |
|  | (0.00856) | (0.00736) | (0.00548) | (0.00526) |
| Observations | 529 227 | 529 227 | 529 227 | 317 252 |

*Note:* 'MAR' refers to medically assisted reproduction, 'SC' stands for spontaneous conception. Standard errors are reported in parentheses. \*\*\* p<0.01, \*\* p<0.05, \* p<0.1.
*Source:* Authors' calculations from Finnish population register data.

**Supplementary Table S5.** Birth outcomes (95% CI) of first-born children in Finland, 1996-2020, by mode of conception and maternal partnership status: a) birth weight; b) gestational age; c) low birth weight; d) preterm. Singletons only.

| Birth outcomes |  |  |
| --- | --- | --- |
|  | Baseline model | Fully adjusted model |
| a) Birth weight (in grams). Coefficients from linear models |  |  |
| SC single | -24 (-47, -1) | -40 (-64, -17) |
| MAR partnered | -20 (-44, 3) | -47 (-70, -23) |
| SC partnered | 30 (7, 53) | -10 (-33, 13) |
| b) Gestational age (in days). Coefficients from linear models |  |  |
| SC single | 1.6 (1.1, 2.2) | 1.1 (0.5, 1.7) |
| MAR partnered | -0.6 (-1.1, 0.0) | -1.0 (-1.5, -0.4) |
| SC partnered | 2.0 (1.5, 2.5) | 1.4 (0.8, 1.9) |
| c) Low birth weight. Odds ratios from logistic models |  |  |
| SC single | 1.06 (0.86, 1.31) | 1.22 (0.99, 1.50) |
| MAR partnered | 1.21 (0.98, 1.48) | 1.43 (1.16, 1.76) |
| SC partnered | 0.78 (0.63, 0.95) | 1.01 (0.83, 1.24) |
| d) Preterm. Odds ratios from logistic models |  |  |
| SC single | 0.81 (0.68, 0.96) | 0.92 (0.77, 1.10) |
| MAR partnered | 1.14 (0.96, 1.36) | 1.25 (1.05, 1.49) |
| SC partnered | 0.70 (0.59, 0.83) | 0.82 (0.69, 0.98) |

*Note:* 'MAR' refers to medically assisted reproduction, 'SC' stands for spontaneous conception. 95% CI are presented in parentheses. Fully adjusted models are controlled for period, child's sex, maternal age, level of education, income, smoking during pregnancy, psychotropic medication use 5 years pre-MAR.
*Source:* Authors' calculations from Finnish population register data.

**Supplementary Table S6.** Proportion of multiple births among of first-born children in Finland, 1996-2020, by mode of conception and maternal partnership status.

|  | 1996-2000 | 2001-2005 | 2006-2010 | 2011-2015 | 2016-2020 |
| --- | --- | --- | --- | --- | --- |
| MAR single | 12.4 | 10.7 | 10.5 | 4.6 | 3.6 |
| SC single | 1.1 | 1.1 | 1.0 | 0.9 | 1.2 |
| MAR partnered | 14.0 | 12.1 | 9.0 | 5.7 | 3.9 |
| SC partnered | 1.0 | 1.1 | 1.1 | 1.1 | 1.1 |

*Note:* 'MAR' refers to medically assisted reproduction, 'SC' stands for spontaneous conception.
*Source:* Authors' calculations from Finnish population register data.

